# Frontal and cerebellar atrophy supports FTSD-ALS clinical continuum

**DOI:** 10.1101/19007831

**Authors:** Beatrice Pizzarotti, Fulvia Palesi, Paolo Vitali, Gloria Castellazzi, Nicoletta Anzalone, Elena Alvisi, Daniele Martinelli, Sara Bernini, Matteo Cotta Ramusino, Mauro Ceroni, Giuseppe Micieli, Elena Sinforiani, Egidio D’Angelo, Alfredo Costa, Claudia AM Gandini Wheeler-Kingshott

## Abstract

**Background:** Frontotemporal Spectrum Disorder (FTSD) and Amyotrophic Lateral Sclerosis (ALS) are neurodegenerative diseases often considered as a continuum from clinical, epidemiologic and genetic perspectives. We used localized brain volume alterations to evaluate common and specific features of FTSD, FTSD-ALS and ALS patients to further understand this clinical continuum.

**Methods:** We used voxel-based morphometry on structural MRI images to localize volume alterations in group comparisons: patients (20 FTSD, seven FTSD-ALS, 18 ALS) versus healthy controls (39 CTR), and patient groups between themselves. We used mean whole-brain cortical thickness 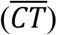 to assess whether its correlations with local brain volume could propose mechanistic explanations of the heterogeneous clinical presentations. We also assessed whether volume reduction can explain cognitive impairment, measured with frontal assessment battery, verbal fluency and semantic fluency.

**Results:** Common (mainly frontal) and specific areas with reduced volume were detected between FTSD, FTSD-ALS and ALS patients, confirming suggestions of a clinical continuum, while at the same time defining morphological specificities for each clinical group (e.g. a difference of cerebral and cerebellar involvement between FTSD and ALS). 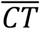values suggested extensive network disruption in the pathological process, with indications of a correlation between cerebral and cerebellar volumes and 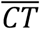 in ALS. The analysis of the neuropsychological scores indeed pointed towards an important role for the cerebellum, along with fronto-temporal areas, in explaining impairment of executive and linguistic functions.

**Conclusions:** We identified common elements that explain the FTSD-ALS clinical continuum, while also identifying specificities of each group, partially explained by different cerebral and cerebellar involvement.

## Introduction

Frontotemporal Spectrum Disorder (FTSD) represents 5% of all causes of dementia in subjects over 65 years and has two main clinical presentations: the behavioral (bvFTSD) and the linguistic variant (Primary Progressive Aphasia, PPA) (Harciarek, 2013). Amyotrophic Lateral Sclerosis (ALS) is a neurodegenerative disease affecting electively the upper and lower motoneuron but several studies have proven that ALS show also cognitive impairment in different domains, like social cognition, verbal memory and executive functions (Leslie *et al*., 2015). Family forms combining both diseases have already been described, hence FTSD and ALS may be thought as pathophysiological continuum, with up to 50% ALS patients presenting FTSD symptoms (Lattante *et al*., 2015). However, it is hard to predict which patients are prone to develop both aspects of the continuum.

Magnetic resonance imaging (MRI) is commonly used to exclude secondary causes of dementia and to detect morphological findings useful for a correct diagnosis. Several voxel based-morphometry (VBM) investigations have used structural MRI images to demonstrate volume alterations in specific areas, such as frontal and temporal lobes, insula and anterior cingulum, in FTSD-ALS continuum. Nevertheless, only a few studies looked at cortical thickness as a marker of the FTSD-ALS continuum and mainly focused on ALS patients (Hendrikse *et al*., 2015; Michael W Weiner An-TAo Du, 2007; Schuster *et al*., 2014).

This study aimed to identify the cognitive and neurostructural deterioration of the FTSD-ALS continuum assessing (i) common and specific areas of volume reductions, as identified with VBM, in ALS, FTSD-ALS and FTSD; (ii) whether whole-brain mean cortical thickness 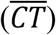 (as surrogate marker of an overall pathological/functional degeneration) could explain differences between groups and propose possible mechanistic interpretations of the different clinical presentations; (iii) whether cognitive and neurostructural deterioration are directly related.

## Materials and methods

### Subjects

Fourty-five patients belonging to FTSD-ALS continuum were recruited at the IRCCS Mondino Foundation from September 2013 to March 2016. Patients underwent a complete diagnostic workup including neuropsychological assessment, MRI and electroneuromyography, in order to obtain an exhaustive phenotypic profiling and a correct etiological definition. Based on the most recent diagnostic criteria, patients were classified into three etiological groups: FTSD (including bvFTSD (Katya Rascovsky, 2013) and PPA(Gorno-Tempini *et al*., 2011)), ALS (Carvalho and Swash, 2009) and FTSD-ALS. According to Rascovsky diagnostic criteria, FTSD diagnosis was supported, but not determined by the cognitive profile. No patient was excluded on the basis of the neuropsychological profile, if diagnostic criteria were still met. As well, ALS diagnosis was made in patients fulfilling Awaji criteria (Costa *et al*., 2012), not excluding subjects with a cognitive impairment, as previously. Patients who met both diagnostic criteria were classified in the FTSD-ALS group (Costa *et al*., 2012; Katya Rascovsky, 2013). During the enrollment, MRI was used with prominent exclusion function of secondary causes of cognitive decline.

A group of 39 age- and sex-matched healthy controls (CTR) were selected as a reference group and enrolled on a voluntary basis among subjects attending a local third-age university (University of Pavia, Information Technology course) or included in a program on healthy aging (Fondazione Golgi, Abbiategrasso). All CTR underwent a clinical assessment to exclude any cognitive or motoneuron impairment.

Exclusion criteria included at least one of the following: major psychiatric disorders, pharmacologically treated delirium or hallucinations, secondary causes of cognitive decline (e.g. vascular, metabolic, endocrine, toxic, iatrogenic).

### Standard Protocol Approvals, Registrations, and Patient Consents

This study was carried out in accordance with the Declaration of Helsinki with written informed consent from all subjects. The protocol was approved by the local ethic committee of the IRCCS Mondino Foundation.

### Neuropsychological assessment

Fourty-three of fourty-five patients underwent a complete neuropsychological evaluation including the following cognitive domains: attention (attentive matrices, trail making test A and B, Stroop test), memory (digit span, verbal span, Corsi block-tapping test, logical memory, Rey–Osterrieth complex figure recall, Rey 15 item test), language (verbal (FAS) and semantic fluency (SF)), executive function (Raven’s matrices, Wisconsin card sorting test, frontal assessment battery (FAB)), and visuo-spatial skills (Rey–Osterrieth complex figure). Cognitive scores were corrected by age and education and compared to the normative cut-off for the Italian population. Two FTSD patients did not undergo the cognitive assessment due to poor collaboration.

FAB, FAS and SF were selected for the correlation analyses as representative of the executive and linguistic functions usually affected in the FTSD-ALS spectrum.

### MRI acquisition

All subjects underwent MRI examination within one month from cognitive assessment. A standardized MRI protocol was carried out on a Siemens Skyra 3T scanner (Siemens, Erlangen, Germany) with a 32 channel head-coil. A 3D T1-weighted (3DT1w) structural MPRAGE sequence was setup according to the Alzheimer’s Disease Neuroimaging Initiative protocol (ADNI2) (Jack *et al*., 2015) with the following parameters: TR = 2300 ms, TE = 2.95 ms, TI = 900 ms, flip angle = 9°, 176 sagittal slices, acquisition matrix = 256×256, in-plane resolution = 1.05×1.05 mm^2^, slice thickness = 1.2 mm, acquisition time = 5.12 minutes. Standard clinical sequences were performed to exclude other pathologies.

### VBM analysis

3DT1w images were converted from DICOM to NIFTI format and segmented in their native space into grey matter (GM), white matter (WM) and cerebrospinal fluid (CSF) using the CAT12 (Gaser and Kurth, 2016) Matlab toolbox for SPM12. The segmented images were modulated and normalized to the Montreal Neurological Institute (MNI) space (ICBM-152 template) with 1.5 mm isotropic voxels, total intracranial volume (TIV) and the mean CT value 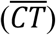 over the whole cortex were assessed with CAT12 (Gaser and Kurth, 2016). The resulting modulated normalized GM and WM images were smoothed using a gaussian kernel of 6×6×6 mm^3^ in SPM12 (John, 2013) and were used as inputs for the statistical analysis.

### Statistical analysis

Demographic and neuropsychologic data were compared using the Statistical Package for the Social Sciences, SPSS21 (IBM, Armonk, New York), to assess significant differences between groups. Gaussian distribution was checked with a Shapiro-Wilk test, then normally distributed variables (age, MMSE and SF) were compared using a one-way ANOVA test with Bonferroni correction, while non-normally distributed ones (FAB and FAS) were compared using a Kruskall-Wallis test (Mann-Whitney for pair comparisons). Categorical variables were compared with a chi-squared test. Two-sided *p*<0.05 was used as significance threshold.

Each group of patients (FTSD, ALS, FTSD-ALS) was compared voxelwise to the CTR group using a one-way ANOVA VBM analysis, performed with SPM12, to identify the atrophic regions of GM and WM specific for each patient group. The same analysis was carried out between pairs of patient groups. In order to assess the potential functional implications of the atrophic areas, we classified all altered voxels based on a >10% spatial overlap between clusters of volume reductions between patients (FTSD, FTSD-ALS, ALS) and CTR and standard resting state networks (RSNs) (Castellazzi *et al*., 2018).

SPM12 was also used to perform multiple regression analyses on all subjects to correlate GM and WM volume with CT values. For each neuropsychological score, a multiple regression analysis was performed on all patients considered together to determine possible areas responsible for the distribution of results.

For all voxelwise analyses, the significance was set at p<0.05 FWE corrected at cluster level. Exploratory results were also investigated with an uncorrected p<0.001 together with a cluster extension correction of minimum 160 voxels. Sex, age and TIV were used as covariates. The XJVIEW toolbox (http://www.alivelearn.net/xjview/) and FSL anatomical atlases, such as JHU (Oishi *et al*., 2008) and SUIT (Diedrichsen *et al*., 2009) were used to accurately localize the regions affected by alterations.

### Data availability statement

Anonymized data of this study will be shared by request from any qualified investigator.

## Results

Overall this study was able to identify specific patterns of volume reduction in ALS, FTSD-ALS and FTSD patients compared to CTR subjects. Whole brain mean CT was found to correlate with GM and WM volumes, non-necessarily implicated in group differences. Correlations of volume and neuropsychological scores in the overall patient group indicated that the cerebellum was a key area for the investigated functions, despite atrophy per se was affecting the cerebellum only in the FTSD group.

### Patient characteristics

Based on clinical criteria, patients were clustered as follows: 20 FTSD (16 bvFTSD and four PPA), 18 ALS and seven patients showing mixed features with FTSD-ALS. Demographic data and cognitive scores are summarized in Table 1.

**Table 1:**
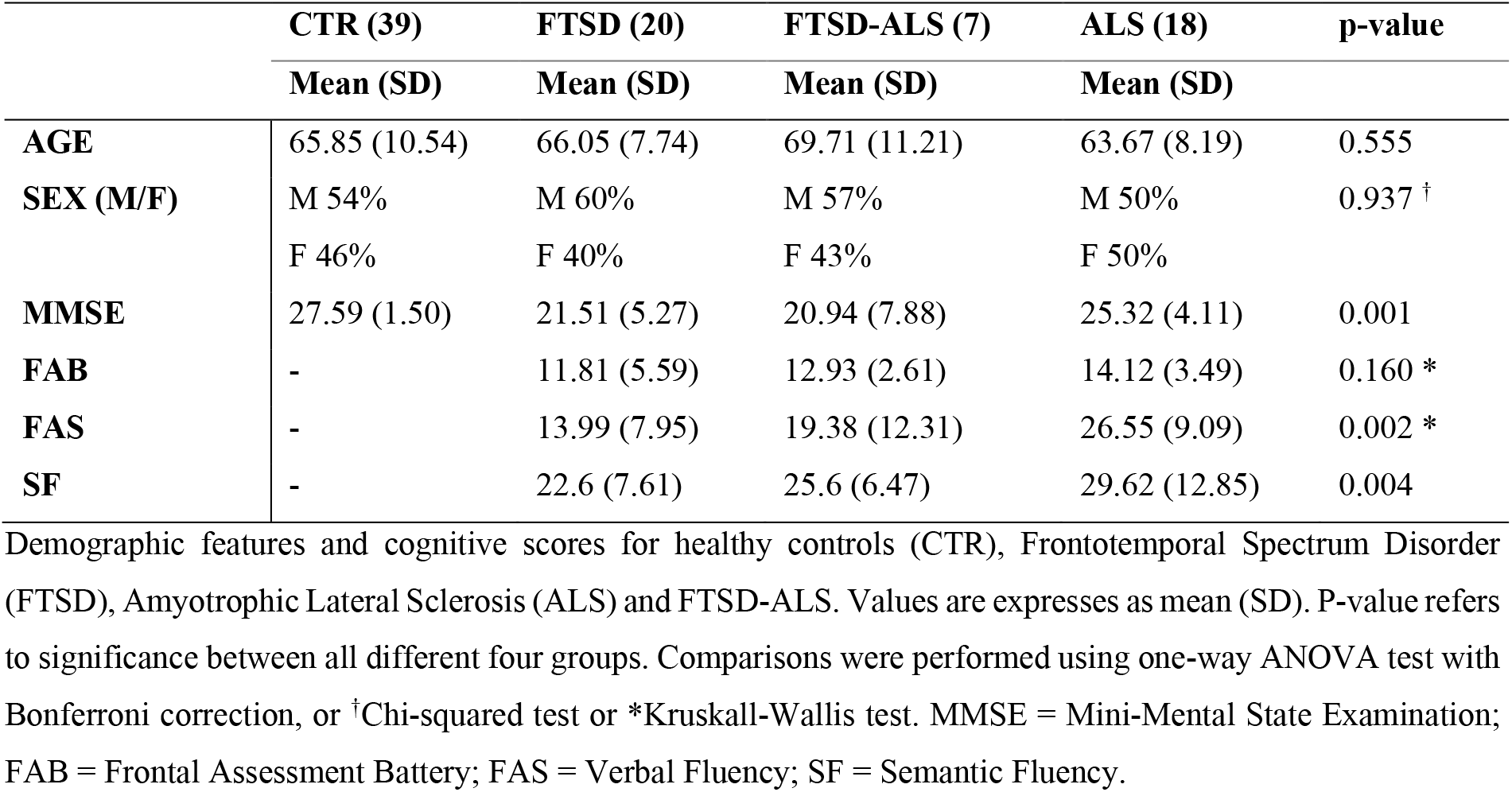
Demographic features and cognitive status.

|  | CTR (39) | FTSD (20) | FTSD-ALS (7) | ALS (18) | p-value |
| --- | --- | --- | --- | --- | --- |
|  | Mean (SD) | Mean (SD) | Mean (SD) | Mean (SD) |  |
| AGE | 65.85 (10.54) | 66.05 (7.74) | 69.71 (11.21) | 63.67 (8.19) | 0.555 |
| SEX (M/F) | M 54%<br>F 46% | M 60%<br>F 40% | M 57%<br>F 43% | M 50%<br>F 50% | 0.937 <sup>†</sup> |
| MMSE | 27.59 (1.50) | 21.51 (5.27) | 20.94 (7.88) | 25.32 (4.11) | 0.001 |
| FAB | - | 11.81 (5.59) | 12.93 (2.61) | 14.12 (3.49) | 0.160 * |
| FAS | - | 13.99 (7.95) | 19.38 (12.31) | 26.55 (9.09) | 0.002 * |
| SF | - | 22.6 (7.61) | 25.6 (6.47) | 29.62 (12.85) | 0.004 |

Demographic features and cognitive scores for healthy controls (CTR), Frontotemporal Spectrum Disorder (FTSD), Amyotrophic Lateral Sclerosis (ALS) and FTSD-ALS. Values are expressed as mean (SD). P-value refers to significance between all different four groups. Comparisons were performed using one-way ANOVA test with Bonferroni correction, or <sup>†</sup>Chi-squared test or \*Kruskal-Wallis test. MMSE = Mini-Mental State Examination; FAB = Frontal Assessment Battery; FAS = Verbal Fluency; SF = Semantic Fluency.

Etiological groups and CTR were age- and sex-matched. MMSE was significantly reduced in FTSD and FTSD-ALS patients with respect to CTR but did not differ (p=0.091) between the three patient groups (ASL, FTSD and FTSD-ALS). FAB scores were homogeneous between patient groups (p=0.160), whereas increasing FAS and SF scores were found from FTSD to ALS group (p=0.002 and p=0.004).

### Comparison between patients and controls

Voxelwise comparisons between patients and CTR with regard to brain volume reductions are reported in Table 2. The most compromised group in terms of GM atrophy is the FTSD group, followed by FTSD-ALS and by ALS. In detail, GM regions with reduced volume in FTSD were mainly located (bilaterally) in the frontal and temporal lobes, while WM regions with reduced volume involved several tracts mainly connecting the frontal and the temporal lobes (Figure 1).

**Table 2:**
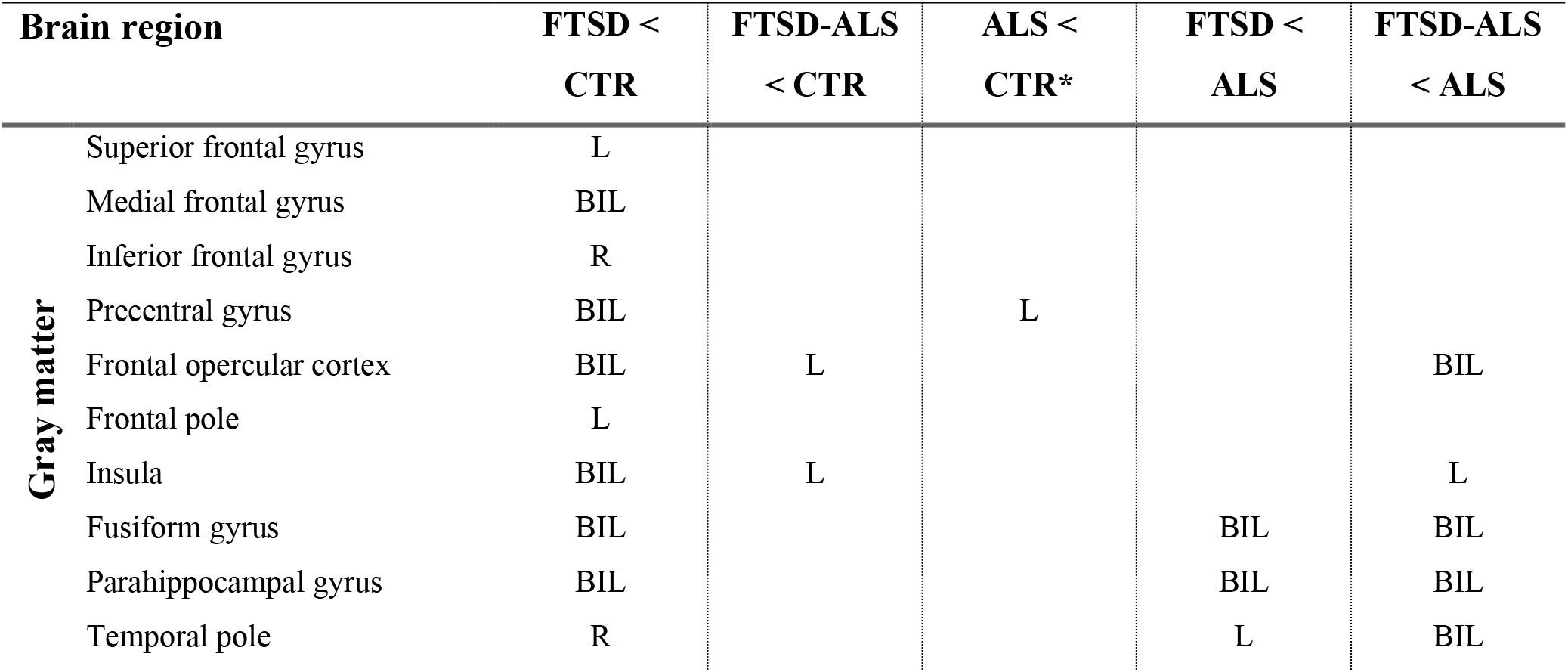
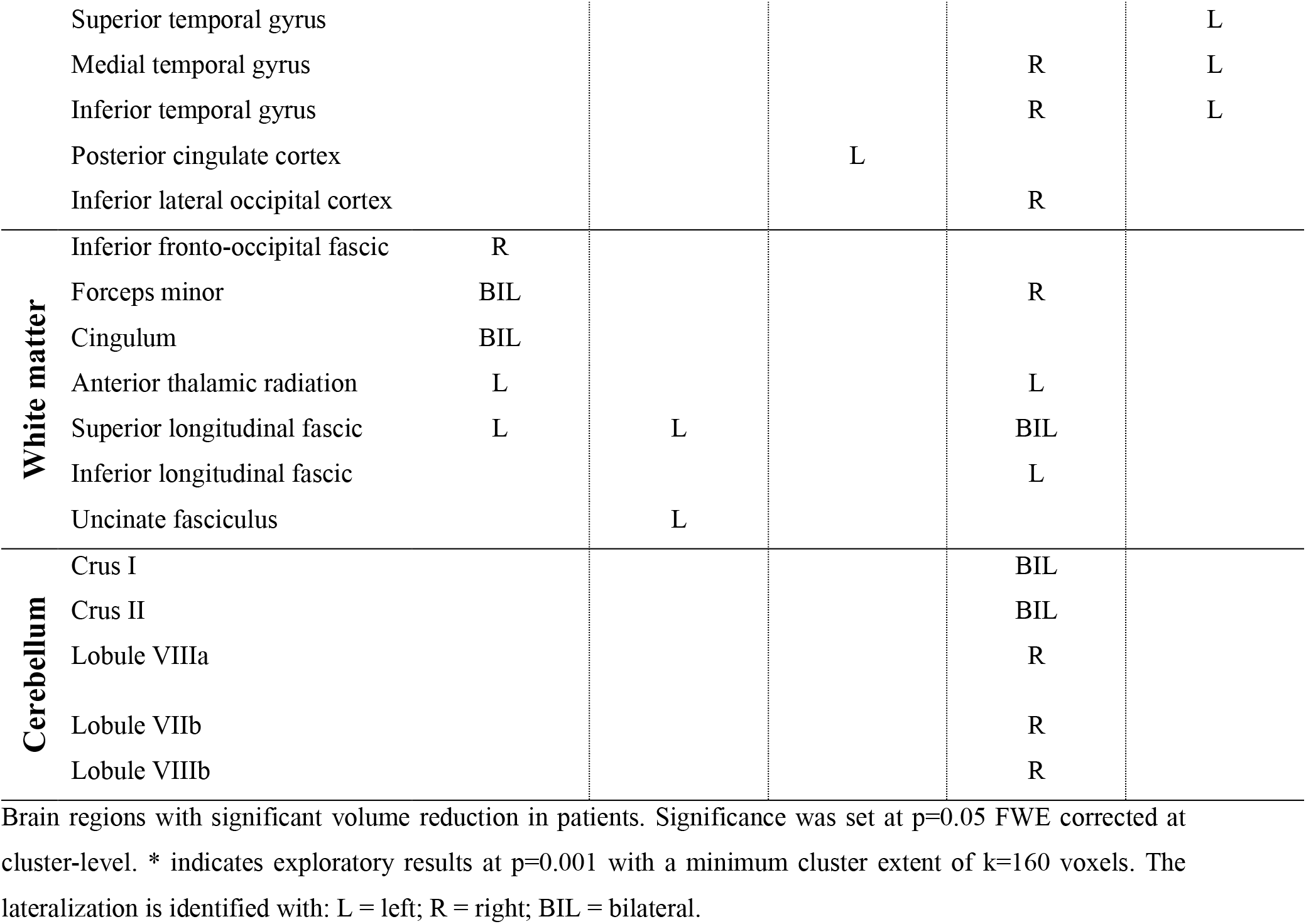
Regions of reduced volume between different groups of patients and controls.

**Figure 1:**
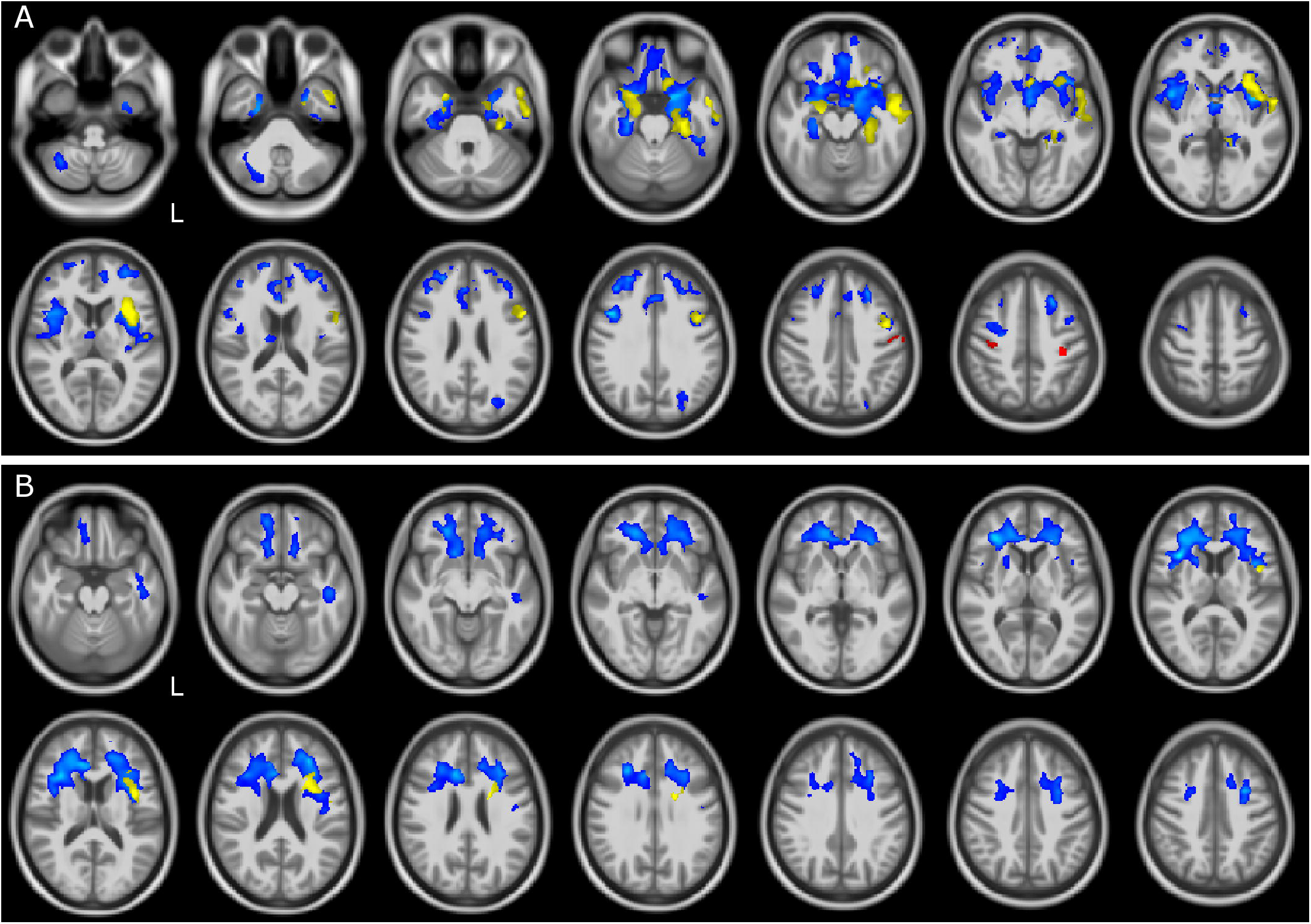
GM and WM volume reduction in patients compared to controls. Regions of grey matter (GM) and white matter (WM) volume reduction in patients compared to controls (CTR). Significance was set at p<0.05 FWE corrected at cluster level, except for the comparison ALS and CTR (p=0.001, k=160). All results are overlaid onto the MNI 152 template and are shown as interleaved axial slices. L indicates the left hemisphere (radiological view). A) GM atrophic regions in FTSD (blue), FTSD-ALS (yellow) and ALS (red) compared to CTRB) WM atrophic regions in FTSD (blue) and FTSD-ALS (yellow) compared to CTR. Blue clusters in A) are showing that compared to CTR, volume reductions are predominant in FTSD compared to the other forms of disease and are distributed across brain regions involving deep GM and cerebral frontal cortex; yellow clusters are showing that FTSD-ALS patients present lesser involvement of degeneration across brain regions while red clusters are identifying mainly motor areas well known to be affected in ALS. For what concerns WM alterations as shown in B), blue clusters are predominant and demonstrate a major involvement of pathways in FTSD, while yellow clusters are fewer and more lateralized in FTDS-ALS. No clusters are emerging as altered in ALS.

Altered regions in FTSD-ALS were lateralized to the left hemisphere and involved GM of the frontal lobe, left insula, and WM of the temporal lobe.

ALS did not show any areas with reduced volume; lowering the statistical threshold, though, a reduced GM volume was found in the left pre- and post-central gyri.

The location of all abnormal voxels with reference to RSN involvement is shown in Table 3. The ALS group involved predominantly areas of the sensory motor network (SMN); FTSD-ALS showed atrophy affecting not only motor functions (e.g. frontal cortex (FCN)), but also other sensory networks (e.g. the occipital visual network (OVN)), several higher-functions, as well as hippocampal areas belonging to the default mode network (DMN); FTSD patients presented atrophy involving almost all functional systems, with a further extensive involvement of the DMN and the cerebellar network.

**Table 3:** Overlap between regions of volume reduction and resting state networks.

| GROUP | MVN | CBL | OVN | SMN | DMN | AN | SN | LVN | LVAN | ECN | WMN | FCN | RVAN |
| --- | --- | --- | --- | --- | --- | --- | --- | --- | --- | --- | --- | --- | --- |
| ALS |  |  |  | × |  | × |  |  |  |  |  |  |  |
| FTSD-ALS |  |  | × |  | × | × |  |  | × | × | × | × |  |
| FTSD | × | × | × | × | × | × | × |  | × | × | × | × | × |

Resting state networks (RSNs) involved by atrophy for each group of patients (rows) compared to controls. “×” indicates areas with reduced volume overlapping with at least 10% of voxels belonging to specific RSNs. MVN = Medial Visual Network; CBLN = Cerebellar Network; OVN = Occipital Visual Network; SMN = Sensory Motor Network; DMN = Default Mode Network; AN = Auditory Network; SN = Salience Network; LVN = Lateral Visual Network; LVAN = Left Ventral Attention Network; ECN = Executive Control Network; WMN = Working memory Network; FCN = Frontal Cortex Network; RVAN = Right Ventral Network.

### Comparison between patient groups

Comparisons between atrophic regions (meaning volume reduction of GM or WM) in different groups of patients are also reported in Table 2. Direct comparison between patient groups showed differences when comparing FTSD and FTSD-ALS to ALS patients (Figure 2). FTSD were more atrophic than ALS in several GM temporal areas, in WM regions connecting the frontal and the temporal lobes (the same involved in the comparison with CTR), and extensively in the posterior cerebellum (Crus I/II, lobules VII and VIII).

**Figure 2:**
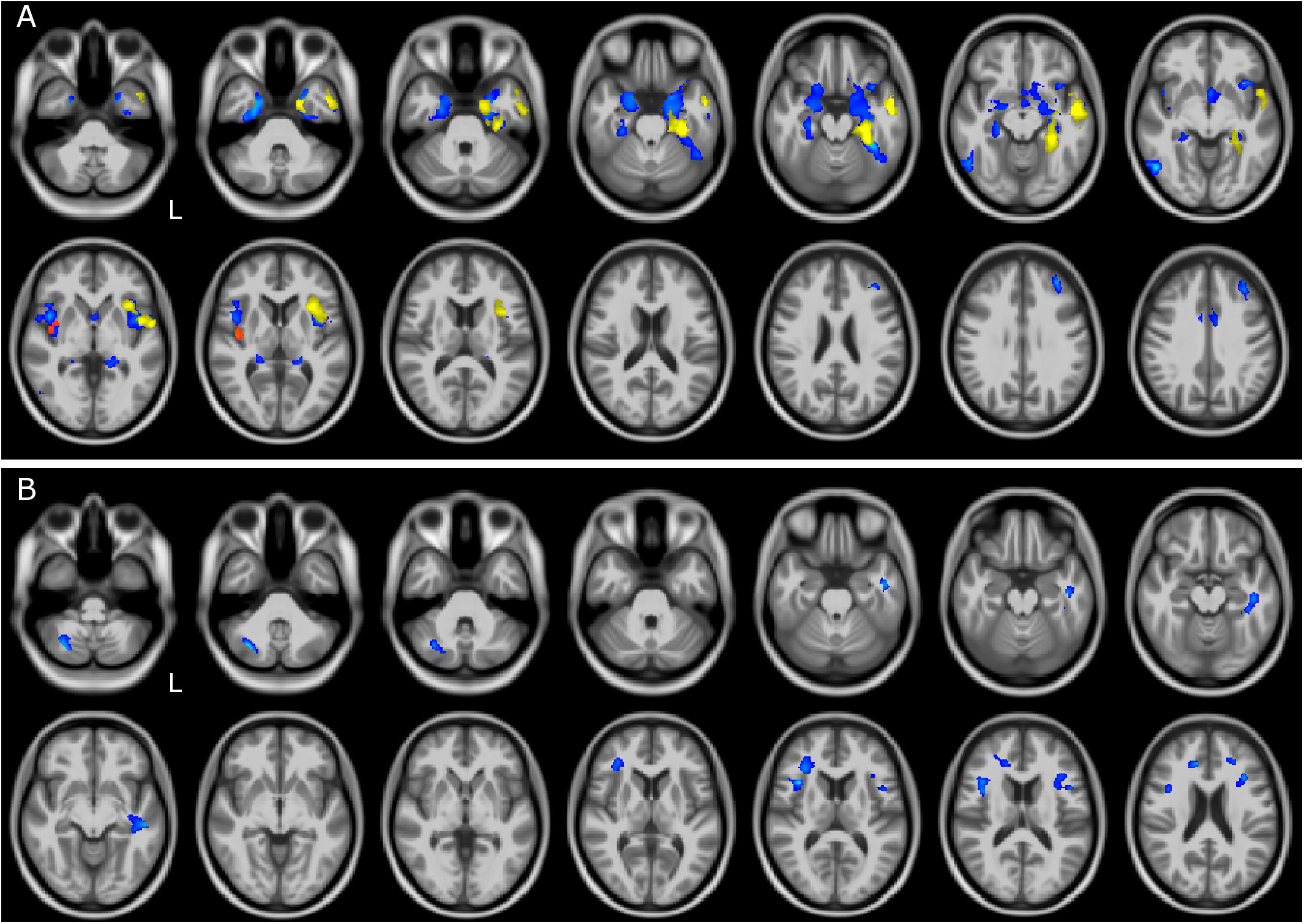
GM and WM volume reduction between patient groups. Regions of grey matter (GM) and white matter (WM) volume reduction between patients. Significance was set at p<0.05 FWE corrected at cluster level, except for the comparison FTSD and FTSD-ALS (p=0.001, k=160). All results are overlaid onto the MNI 152 template and are shown as interleaved axial slices. L indicates the left hemisphere (radiological view). A) GM atrophic regions in FTSD vs ALS (blue), FTSD-ALS vs ALS (yellow) and FTSD vs FTSD-ALS (red). B) WM atrophic regions in FTSD (blue) compared to ALS. Differences between groups are demonstrating both GM and WM volume reductions in FTSD compared to ALS, as shown by blue clusters in A) and B). There are also a number of yellow clusters in A) identifying more severe GM volume reductions in FTSD-ALS compared to ALS patients, while no differences were detected in WM between these two groups as shown by the lack of yellow clusters in B). Differences in terms of volume reduction between FTSD and FTSD-ALS were localized to a lateralized cluster of GM as shown in A).

FTSD-ALS in comparison to ALS shared several areas of GM atrophy that emerged in the comparison of FTSD to ALS. These areas involved mainly GM of the temporal lobe.

Comparisons of FTSD versus FTSD-ALS did not survive FWE correction. Lowering the statistical threshold, it emerged that in FTSD the insula is the only area with reduced volume than FTSD-ALS. No WM regions seemed to indicate group specific pattern of volume loss between FTSD and FTSD-ALS and between FTSD-ALS and ALS.

### Correlation between volume and CT

Correlations between volume and whole brain CT in CTR, all patients and different patient groups are reported in the supplementary material.

In CTR, positive correlation was found between CT and volume of several GM regions of the frontal, parietal and temporal lobes. In the ALS group, lower CT correlated with lower GM volume in the cerebellum, while lowering the statistical threshold showed that also WM regions connecting frontal and temporal lobes are involved.

### Correlation between volume and neuropsychological scores

Correlations between neuropsychological scores and volume in all patients are reported in Table 4. Lower FAB scores correlated with lower GM volume in several cerebellar areas, while no correlations were found in WM regions.

**Table 4:** Neuropsychological regression analysis.

| Brain region |  | FAB | FAS | SF |
| --- | --- | --- | --- | --- |
| **White matter** | SLF (frontal/temporal part) |  | R |  |
|  | SLF (parietal part) |  | L |  |
|  | Anterior thalamic radiation |  | L |  |
| **Cerebellum** | Lobule IV |  | BIL |  |
|  | Lobule V |  | R |  |
|  | Lobule VI |  | BIL |  |
|  | Crus I | L | BIL |  |
|  | Crus II | L | BIL |  |
|  | Lobule VII | L | BIL |  |
|  | Lobule VIII | L | BIL |  |
|  | Lobule IX |  | L |  |

Areas of positive correlation between neuropsychological scores and brain volume. Significance was set at $p=0.05$ FWE corrected at cluster-level. The lateralization is identified with: L = left; R = right; BIL = bilateral. SLF = superior longitudinal fasciculus.

Reduced FAS scores had a significant involvement of GM regions of cerebellum, and of WM volume of multiples subcortical regions.

No correlations were found between SF scores and GM or WM volumes.

## Discussion

The main finding of this study supports the clinical continuum of FTSD, FTSD-ALS and ALS patients given the presence of shared common structural and cognitive features, but also specific structural features were identified for each etiological group supporting the fact that the clinical continuum is indeed characterized by three different diagnostic entities. The clinical continuum was well detected by fluency scores (both FAS and SF), which were the lowest in FTSD, lower in FTSD-ALS and only slight decreased in ALS with respect to normal scores. The same behavior was detected in volume deterioration: FTSD presented a diffuse cerebral (bilateral frontotemporal) and cerebellar atrophy, FTSD-ALS presented a less pronounced cerebral (left frontotemporal) and cerebellar atrophy, while ALS presented a minimal atrophy (bilateral pericentral).

Interestingly, however, there are clear specificities showing involvement of cognitive areas and of WM disruption that contribute to differentiate clinical and neuropsychological presentations. Common features included more atrophic frontal lobes compared to CTR. It is noteworthy that while areas of GM atrophy were found in all three groups of patients compared to CTR, WM atrophy was more disease specific, with extensive involvement in FTSD and some involvement in FTSD-ALS.

Atrophy of frontal and temporal cortices in FTSD patients confirms previous results (Kanda *et al*., 2008), whereas spread reduced WM volumes indicates an overall network disruption that may be independent or secondary to GM atrophy. The present cross-sectional data cannot answer mechanistic questions on WM and GM alterations in FTSD patients, that need to be dealt with appropriate dedicated longitudinal studies where the interplay of GM and WM involvement can be followed over time.

FTSD-ALS patients, instead, showed lateralized alterations (to the left hemisphere) in the same frontal and temporal GM areas that are also involved in FTSD. Given that the insula has a pivotal role in cognitive functions (self-perception, motivation, executive functions and subjective responses) and the anterior insula is connected with dorsolateral and ventromedial prefrontal cortex (Namkung *et al*., 2017) it is interesting that this brain region is more and bilaterally atrophic in the FTSD group with worse executive functions. The FTSD-ALS group showed involvement of some WM regions belonging to the superior longitudinal fasciculus (SLF), also altered in FTSD, as well as of the uncinate fasciculus (UF). The SLF and UF are both associative long tracts that connect different lobes of the brain, with the SLF being known to contribute to higher motor functions while the UF has a role in memory and emotional behavior (Heide *et al*., 2013). This finding supports the mixed clinical presentation of FTSD-ALS patients.

In ALS patients, previous studies reported atrophy in non-motor areas involved in executive and behavioral functions, such as frontal, temporal and limbic regions (Menke *et al*., 2014). Although our ALS patients did not show atrophy in those regions, the involvement of motor and premotor regions emerging from a less stringent statistical analysis is indeed consistent with motor symptoms onset in ALS.

These results were also captured by the RSN overlap analysis. The ALS group showed an involvement of the sensory motor network (SMN) only, while FTSD-ALS had atrophy spread across sensory and associative networks, including the DMN, although limited to the hippocampus. Intriguing is the lack of volume reduction in motor areas in the FTSD-ALS group, which is captured also by the absence of SMN involvement. This is probably due to the statistical power of the VBM analysis as the precentral gyrus is different between FTSD-ALS and CTR, but does not survive multiple comparisons. In FTSD there was a widespread involvement not only of sensory and associative networks, but of all cognitive domains including executive function networks. These three groups of patients can be considered as a clinical continuum, where subjects belong to one group or the other depending on the domain affected by tissue atrophy.

Furthermore, the direct comparison between patients highlighted that some regions of the temporal lobe had reduced volume in FTSD and FTSD-ALS compared to ALS. Interestingly, FTSD showed reduced cerebellar volume compared to ALS, which confirms findings of previous studies in C9orf72 mutated patients (Tan *et al*., 2014). Genetic data were not available for our analysis, but it would be interesting to understand whether cerebellar involvement is gene-dependent. Moreover, since our FTSD group was mainly represented by the behavioral variant (16 subjects), we could also hypothesize that cerebellar alterations, which were shown in this group, are particularly relevant to this disease phenotype. The fact that cerebellar Crus I/II (bilaterally) was involved in FTSD compared to ALS, could partially explain the cognitive impairment of these patients given the cerebellar role in memory and language processing (Gellersen *et al*., 2017) as well in continuous cognitive processing tasks (Castellazzi *et al*., 2018). Furthermore, our study shows an involvement of the posterior cerebellum in FTSD compared to ALS, which could point to a greater disruption of the cerebro-cerebellar circuit in FTSD. This is further supported by the reduced volume of the anterior thalamic radiation, which is known to be part of the efferent pathway from the superior cerebellar peduncle (Palesi *et al*., 2015).

In order to understand the source of atrophy in the three patient groups and help mechanistic interpretation of the VBM results, we investigated the correlation between 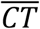 values and volume, as measured by VBM analysis. Indeed, both VBM volume changes and cortical thickness measurements are based on cortex morphology, but with cortical thickness being more specific to cellular density. Details of 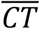 correlations with local volumes are given in the supplementary materials; nevertheless, it is worth noting that the correlation between 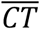 and local volume of long WM bundles in ALS, could suggest a key role of inter-lobe WM integrity for cognitive functions. Alterations of 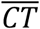 and temporal WM in FTSD, instead, is consistent with emotional and cognitive impairment in this group of patients.

The correlation between neuropsychological scores and brain volume was performed to elucidate whether the cognitive involvement could be described in terms of atrophy of specific brain regions. The correlation between neuropsychological scores and brain volume for the overall patient group is consistent with recent literature showing more and more often that the cerebellum has a key role in cognition and in supporting advanced functions (Castellazzi *et al*., 2018). Furthermore, recent studies have reported the presence of a high proportion of cerebellar connections with the frontal and prefrontal cortex (Palesi *et al*., 2017) consistent with the fact that the FAB is thought to require predominantly frontal and prefrontal cortex and more generally high-level functions. Indeed, Crus I is known to be involved in cognition, whereas lobule VII has recently been shown to have a role in cognitive and social behavior, with particular focus on persisting behavior and novelty seeking (Badura *et al*., 2018). Since the cerebellar areas correlating with FAB are also those resulting more atrophic in FTSD compared to ALS (i.e. Crus I/II and lobule VII/VIII), it is possible that the correlation between cerebellar volume and neuropsychological scores is driven by alterations of the FTSD group. Future studies will be able to confirm the generalization of these results for the FAB test. Our findings also revealed that lower performances of the verbal fluency test, i.e. FAS, correlated with reduced volume of both frontal areas, consistently with their inhibitory role, and with mostly bilateral cerebellar areas, including Crus I/II as well as both the anterior (lobule IV, V, VI) and posterior (lobule VII, VIII and IX) cerebellum. The extensive cerebellar involvement can be explained by the amnestic and linguistic roles of Crus I/II and by the motor involvement of the anterior cerebellum (Gellersen *et al*., 2017).

These interesting results, however, must be interpreted with caution. The relatively small number of patients per group, in particular for FTSD-ALS, may have reduced the statistical power of some analysis; moreover, within groups there were possible sub-groups like in FTSD where both linguistic and behavioral variants were included together, potentially reducing sensitivity to detect further significant differences. Nonetheless, it is important to consider that FTSD and ALS are rare diseases so larger cohorts may be feasible in future multi-center studies. Finally, the CTR group did not undergo the neuropsychological examination, therefore limiting the correlation analysis to patients. Having CTR scores would be highly desirable for future studies.

In conclusion, our study assessed morphological alterations of FTSD, FTSD-ALS and ALS patients in the attempt to clarify the substrate of known clinical differences and their clinical continuum. The involvement of GM areas, to different extent, in frontal regions in all groups, with atrophy of insular areas in FTSD and FTSD-ALS patients, and temporal cortices and WM regions in FTSD only, supports the presence of shared features, but, at the same time, very distinctive characteristics of these patient groups. Interestingly, cerebellar differences between FTSD and ALS as well as the cerebellar role in correlations between volume and cognitive scores, indicates that the cerebellum contributes to determining the FTSD or ALS variant of this continuum. Future longitudinal multi-center studies are needed to better investigate the relation between localized volume reduction, clinical and neuropsychological outcomes, and 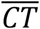 in terms of mechanisms of the FTSD and ALS spectrum.

## Data Availability

Statistical summary maps are available upon request.

## Abbreviations

FTSD: Frontotemporal Spectrum Disorder
ALS: Amyotrophic Lateral Sclerosis
CTR: healthy controls
VBM: voxel based-morphometry
CT: cortical thickness
FAB: frontal assessment battery
FAS: verbal fluency
SF: semantic fluency
GM: grey matter
WM: white matter
TIV: total intracranial volume

## Acknowledgments

We thank the patients, their families, all healthy volunteers for making this research possible. We thank Giancarlo Germani for MRI acquisitions and Roberta Fortunato for her support to the study organization.

## Supplementary material: VBM and 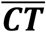

### Methods

SPM12 was used to perform multiple regression analyses on all subjects to correlate GM and WM volume with 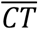 values. For each neuropsychological score, a multiple regression analysis was performed on all patients considered together to determine possible areas responsible for the distribution of results.

### Results

#### Correlation between volume and 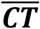

Correlations between volume and whole brain 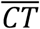 in CTR, all patients and different patient groups are reported in the supplementary material table.

In CTR, positive correlation was found between 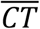 and volume of several GM regions of the frontal, parietal and temporal lobes.

Considering all patients, lower 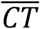 correlated with lower GM volume of areas in the frontal, parietal and temporal lobes, and with WM volume of regions connecting frontal and temporal/occipital lobes.

In the FTSD group, lower 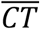 correlated with lower GM volume of areas in the parietal lobe, whereas correlation with volume of WM regions connecting frontal and temporal/occipital lobes emerged lowering the statistical threshold.

In the ALS group, lower 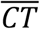 correlated with lower GM volume in the cerebellum, while lowering the statistical threshold also WM regions connecting frontal and temporal lobes are involved.

**Table S1:**
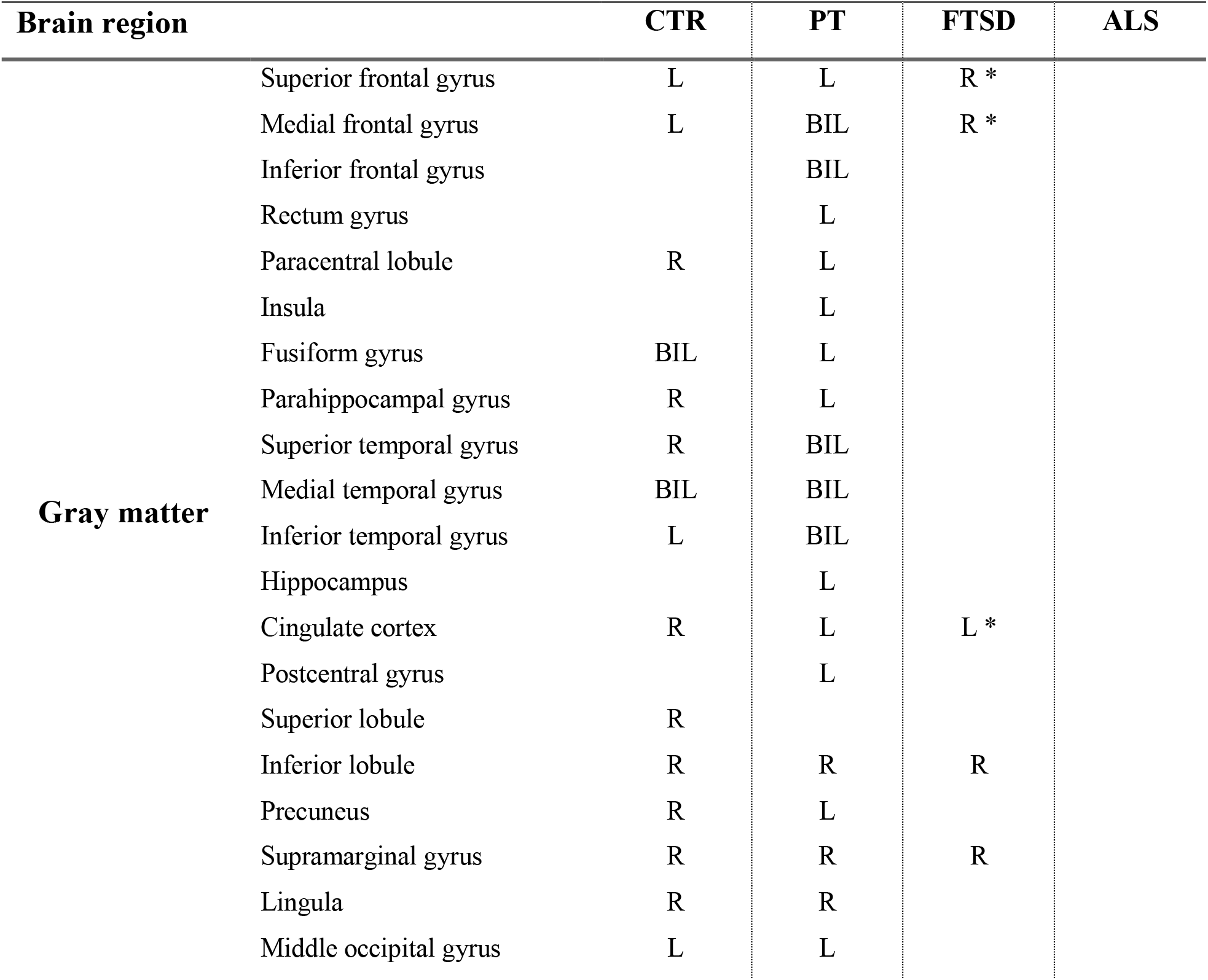
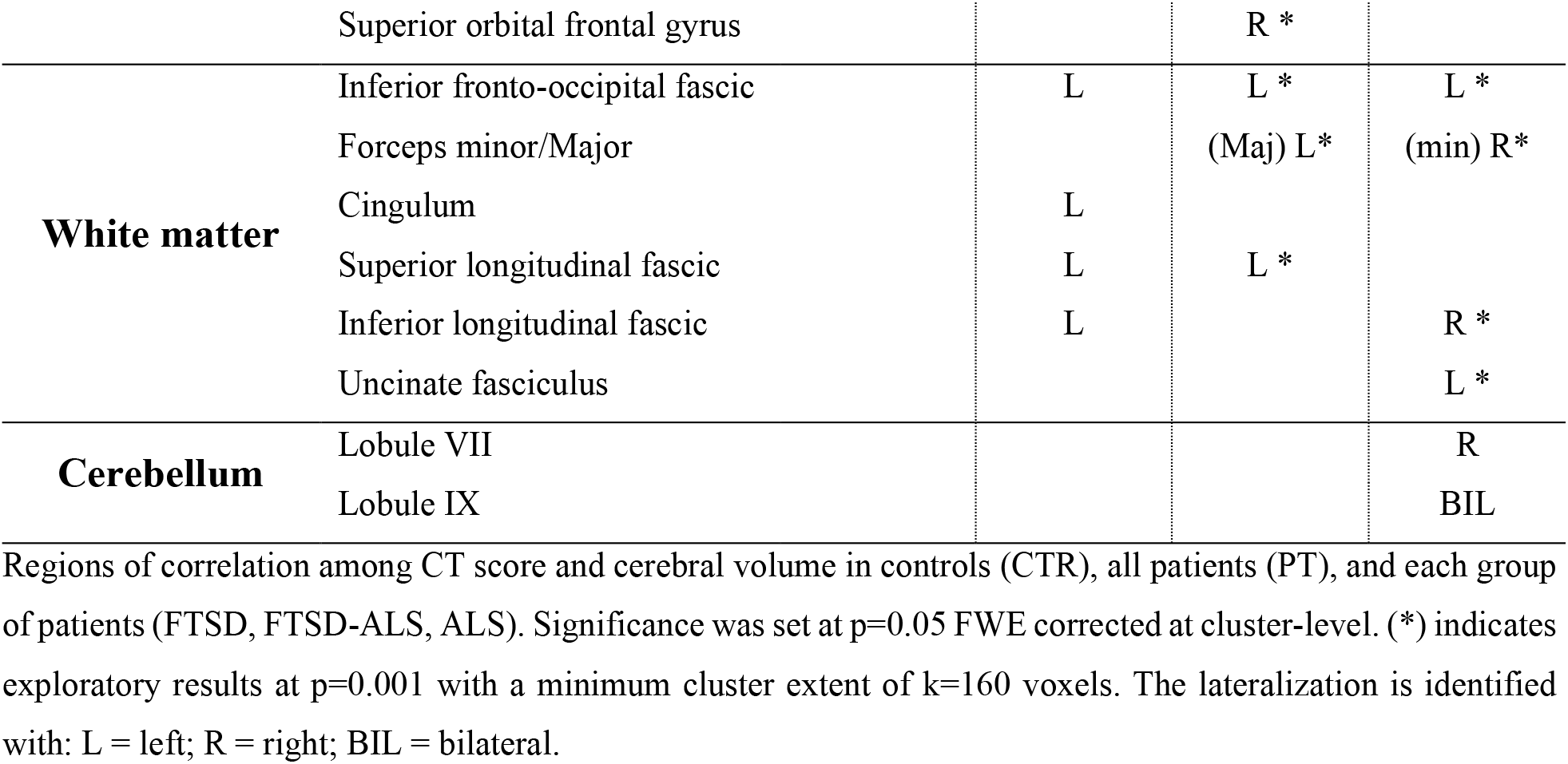
Correlation of local voxel brain morphometry (VBM) properties with whole brain cortical thickness 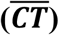.

## Discussion

These are exploratory results (p<0.001, uncorrected) except for the results in the ALS group, it is intriguing to consider 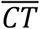 correlations with GM and WM volume in each group. Indeed, the different etiology of these patients brings out some differences in morphological changes that could subtend 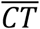 and VBM volumes correlations. In particular, in ALS, 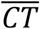 correlates with volume in the posterior cerebellum, and in particular area VIII and IX, known to be key to motor control, as well as motor learning and sensory integration. This is different from previous studies that showed 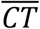 correlations with the precentral cortex, cingulum and insula.^31,32^ In our cohort, though, 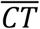 correlated with extensive WM areas affecting long tracts connecting main cortical lobes, including the forceps minor connecting interhemispheric frontal cortices, the inferior longitudinal fasciculus connecting temporal and occipital lobes, the UF connecting the limbic system to the temporal and frontal lobes and the inferior fronto-occipital fasciculus connecting occipital and frontal cortices. These correlations that emerge only in ALS patients, suggest that white matter integrity has a key role in preserving cortical cell density, as measured through 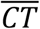. Indeed, no correlations were found in FTSD-ALS, while CT and volume correlated in a number of frontal lobe GM regions and in only temporal WM in FTSD, supporting the different clinical presentation of these patients’ groups.

## References

Badura A, Verpeut JL, Wang SS-H. Normal cognitive and social development require posterior cerebellar activity. eLIFE 2018: 1–36.

Carvalho MD, Swash M. Awaji diagnostic algorithm increases sensitivity of El Escorial criteria for ALS diagnosis. Amyotroph Lateral Scler 2009; 10: 53–57.

Cash DM, Bocchetta M, Thomas DL, Dick KM, Van Swieten JC, Borroni B, et al. Patterns of gray matter atrophy in genetic frontotemporal dementia: results from the GENFI study. Neurobiology of Aging 2018; 62: 191–196.

Castellazzi G, Bruno SD, Toosy AT, Casiraghi L, Palesi F, Savini G, et al. Prominent Changes in Cerebro-Cerebellar Functional Connectivity During Continuous Cognitive Processing. Front. Cell. Neurosci. 2018; 12: 2959–15.

Costa J, Swash M, de Carvalho M. Awaji criteria for the diagnosis of amyotrophic lateral sclerosis:a systematic review. Arch Neurol 2012; 69: 1410–1416.

Christidi F, Karavasilis E, Riederer F, Zalonis I, Ferentinos P, Velonakis G, et al. Gray matter and white matter changes in non-demented amyotrophic lateral sclerosis patients with or without cognitive impairment: A combined voxel-based morphometry and tract-based spatial statistics whole-brain analysis. Brain Imaging and Behavior 2017: 1–17.

Cosottini M, Pesaresi I, Piazza S, Diciotti S, Cecchi P, Fabbri S, et al. Structural and functional evaluation of cortical motor areas in Amyotrophic Lateral Sclerosis. Experimental Neurology 2012; 234: 169–180.

Crespi C, Dodich A, Cappa SF, Canessa N, Iannaccone S, Corbo M, et al. Multimodal MRI quantification of the common neurostructural bases within the FTD-ALS continuum. Neurobiology of Aging 2017: 1–35.

de Carvalho M, Dengler R, Eisen A, England JD, Kaji R, Kimura J, et al. Electrodiagnostic criteria for diagnosis of ALS. Clin Neurophysiol 2008; 119: 497–503.

Diedrichsen J, Balsters JH, Flavell J, Cussans E, Ramnani N. A probabilistic MR atlas of the human cerebellum. NeuroImage 2009; 46: 39–46.

Gaser C, Kurth F. Manual Computational Anatomy Toolbox - CAT12. 2016.

Gellersen HM, Guo CC, O’Callaghan C, Tan RH, Sami S, Hornberger M. Cerebellar atrophy in neurodegeneration-a meta-analysis. [Internet]. Journal of Neurology, Neurosurgery & Psychiatry 2017; 88: 780–788.

Gordon PH. Amyotrophic Lateral Sclerosis: An update for 2013 Clinical Features, Pathophysiology, Management and Therapeutic Trials. A&D 2013; 4: 295–310.

Gorno-Tempini ML, Rascovsky K, Knopman DS, Boeve BF, Cappa SF, Hodges JR, et al. Classification of primary progressive aphasia and its variants. Neurology 2011; 02

Harciarek SCM. Language, Executive Function and Social Cognition in the Diagnosis of Frontotemporal Dementia Syndromes. Int Rev Psychiatry 2013; 25: 178–196.

Heide Von Der RJ, Skipper LM, Klobusicky E, Olson IR. Dissecting the uncinate fasciculus: disorders, controversies and a hypothesis. Brain 2013; 136: 1692–1707.

Hendrikse J, Van Den Berg LH, Walhout R, Westeneng HJ, Van De Heuvel MP, Veldnick JH, et al. Cortical thickness in ALS: towards a marker for upper motor neuron involvement. J Neurol Neurosurg Psychiatry 2015; 86: 288–294.

Jack CR, Barnes J, Bernstein MA, Borowski BJ, Brewer J, Clegg S, et al. Magnetic resonance imaging in Alzheimer’s Disease Neuroimaging Initiative 2. Alzheimers Dement 2015; 11: 740–756.

John A. Generative Models for MRI/DWI. Frontiers in Neuroinformatics 2013; 7

Kanda T, Ishii K, Uemura T, Miyamoto N, Yoshikawa T, Kono AK, et al. Comparison of grey matter and metabolic reductions in frontotemporal dementia using FDG-PET and voxel-based morphometric MR studies. Eur J Nucl Med Mol Imaging 2008; 35: 2227–2234.

Katya Rascovsky MG. Clinical diagnostic criteria and classification controversies in frontotemporal lobar degeneration. Int Rev Psychiatry 2013; 25: 145–158.

Lattante S, Ciura S, Rouleau GA, Kabashi E. Defining the genetic connection linking amyotrophic lateral sclerosis (ALS) with frontotemporal dementia (FTD). Trends Genet. 2015; 31: 263–273.

Leslie FVC, Hsieh S, Caga J, Savage SA, Mioshi E, Hornberger M, et al. Semantic deficits in amyotrophic lateral sclerosis. Amyotrophic Lateral Sclerosis and Frontotemporal Degeneration 2015; 16: 46–53.

Meeter LH, Kaat LD, Rohrer JD, Van Swieten JC. Imaging and fluid biomarkers in frontotemporal dementia. Nat Rev Neurol 2017; 13: 406–419.

Menke RAL, Körner S, Filippini N, Douaud G, Knight S, Talbot K, et al. Widespread grey matter pathology dominates the longitudinal cerebral MRI and clinical landscape of amyotrophic lateral sclerosis. Brain 2014; 137: 2546–2555.

Michael W Weiner An-TAo Du NSJHKHJRMLG-TKRBLM. Different regional patterns of cortical thinning in Alzheinr’s disease and frontotemporal dementia. 2007; 130: 1150–1166.

Namkung H, Kim S-H, Sawa A. The Insula: An Underestimated Brain Area in Clinical Neuroscience, Psychiatry, and Neurology. Trends in Neurosciences 2017; 40: 200–207.

Oishi K, Zilles K, Amunts K, Faria A, Jiang H, Li X, et al. Human brain white matter atlas: identification and assignment of common anatomical structures in superficial white matter. NeuroImage 2008; 43: 447–457.

Palesi F, Rinaldis A, Castellazzi G, Calamante F, Muhlert N, Chard D, et al. Contralateral cortico-ponto-cerebellar pathways reconstruction in humans in vivo: implications for reciprocal cerebro-cerebellar structural connectivity in motor and non-motor areas. Scientific Reports 2017: 1–13.

Palesi F, Tournier JD, Calamante F, Muhlert N, Castellazzi G, Chard D, et al. Contralateral cerebello-thalamo-cortical pathways with prominent involvement of associative areas in humans in vivo. Brain Struct Funct 2015; 220: 3369–3384.

Schuster C, Kasper E, Dyrba M, Machts J, Bittner D, Kaufmann J, et al. Cortical thinning and its relation to cognition in amyotrophic lateral sclerosis. Neurobiology of Aging 2014; 35: 240–246.

Shen D, Cui L, Fang J, Cui B, Li D, Tai H. Voxel-Wise Meta-Analysis of Gray Matter Changes in Amyotrophic Lateral Sclerosis. Front. Aging Neurosci. 2016; 8: 1507.

Tan RH, Devenney E, Dobson-Stone C, Kwok JB, Hodges JR, Kiernan MC, et al. Cerebellar integrity in the amyotrophic lateral sclerosis-frontotemporal dementia continuum. PLoS ONE 2014; 9: e105632.

